# GenomeDiver: A platform for phenotype-guided medical genomic diagnosis

**DOI:** 10.1101/2020.11.22.20236430

**Authors:** Nathaniel Pearson, Christian Stolte, Kevin Shi, Faygel Beren, Noura S. Abul-Husn, Gabrielle Bertier, Kaitlyn Brown, Jacqueline A. Odgis, Sabrina A. Suckiel, Carol R. Horowitz, Melissa Wasserstein, Bruce D. Gelb, Eimear E. Kenny, Charles Gagnon, Vaidehi Jobanputra, Toby Bloom, John M. Greally

## Abstract

**Purpose:** Making a diagnosis from clinical genomic sequencing requires well-structured phenotypic data to guide genotype interpretation. A patient’s phenotypic features can be documented using the Human Phenotype Ontology (HPO), generating terms used to prioritize genes potentially causing the patient’s disease. We have developed GenomeDiver to provide a user interface for clinicians that allows more effective collaboration with the clinical diagnostic laboratory, with the goal of improving the success of the diagnostic process.

**Methods:** GenomeDiver is designed to prompt reverse phenotyping of patients undergoing genetic testing, enriching the amount and quality of structured phenotype data for the diagnostic laboratory, and helping clinicians to explore and flag diseases potentially causing their patient’s presentation.

**Results:** We show how GenomeDiver communicates the clinician’s informed insights to the diagnostic lab in the form of HPO terms for interpretation of genomic sequencing data. We describe our user-driven design process, the engineering of the software for efficiency, security and portability, and an example of the performance of GenomeDiver using simulated genomic testing data.

**Conclusions:** GenomeDiver is a first step in a new approach to genomic diagnostics that enhances laboratory-clinician interactions, with the goal of directly engaging clinicians to improve the outcome of genomic diagnostic testing.

## INTRODUCTION

Compelling economic evidence now supports how genomic sequencing saves both money and time^1,2^ and improves quality-adjusted life years^3^ in the diagnosis of the 3.5–5.9% of the population with rare diseases.^4^ To make this genomic diagnostic process more efficient, substantial attention has been appropriately paid to detecting and understanding the effects of sequence variants in the human genome.^5^ Linking genetic variants to diseases is challenging and is most successful in case-control studies or when comparing multiple affected and unaffected family members, for which excellent analytical approaches have been developed.^6,7^ In a clinical context, however, testing can only be ordered on the proband and one or both parents (if available), with results needed sufficiently rapidly to influence care. The genomic diagnostic laboratory relies on linking the variants present in the patient’s genome with the patient’s phenotypic features. When a pathogenic genetic variant has previously been found to be the cause of a disease, that associated disease is characterized by a set of Human Phenotype Ontology (HPO) terms.^8^ If the patient has a variant that appears potentially pathogenic, causality is supported if the list of HPO terms in the disease associated with a pathogenic variant of that gene significantly overlaps the HPO terms used to describe the patient. A number of phenotype-based variant prediction tools exist to perform this kind of variant prioritization.^9^

If the diagnostic laboratory is provided with a greater number of HPO terms describing the patient on whom sequencing was performed, the diagnostic yield is increased.^10,11^ The comprehensive collection of these HPO terms is very difficult to scale effectively. Ordering genomic tests for rare diseases involves providing the lab with a primary indication for the testing (*e*.*g*. hearing loss, seizures), but also requires communicating additional findings in the patient, sometimes through checklists completed as part of the test order, also potentially involving the diagnostic laboratory reviewing notes from the patient’s health record. The diagnostic laboratory staff then extract information from these sources and can make decisions about how the phenotype can be represented as HPO terms. In critically ill children, an approach has been described that involves automated extraction of HPO terms from the electronic health record (EHR) using natural language processing,^12^ helping with the speed of generation of HPO terms.

We have developed GenomeDiver to help clinicians contribute more effectively to the diagnostic process. GenomeDiver facilitates the curation by the clinician of case-relevant HPO terms quickly, in greater numbers and more accurately than is currently typical, with the goal of improving our ability to make diagnoses using genomic information.

## MATERIALS AND METHODS

### The user-centered design process for GenomeDiver

We developed GenomeDiver using evidence-based participatory methods from design thinking.^13^ User research began with initial interviews with physicians and genetic testing laboratory staff, followed by a one-day stakeholder workshop and a design sprint to understand user needs, to define requirements, to sketch candidate workflows, to choose among them and to prototype. We included physicians (with and without specialist genetics training), genetic counselors, genetic testing laboratory personnel, research scientists, software tool builders, bioinformaticians and biologists, a designer, and a software engineer. The resulting concepts, sketches, and paper prototypes informed the designs for user experience, workflow, and user interfaces.

### The GenomeDiver workflow: overview of a ‘dive’

The workflow for a case (**Figure S1**) begins with the upload by the genetic testing lab of the patient’s genomic information as a variant call format (VCF) file, along with the HPO terms that they derived from the test requisition documents. The clinician then starts a ‘dive’ by selecting the patient from the GenomeDiver interface. The next step, refining the patient’s phenotype, requires the clinician categorize HPO terms generated by GenomeDiver by dragging each into one of three screen areas: ‘Present’ in the patient, ‘Absent’, or ‘Unknown’. Using this updated and enriched phenotypic information, GenomeDiver re-analyzes the genomic data and presents candidate diseases for the clinician to review and flag, adding any comments that they would like to communicate to the diagnostic laboratory. To conclude the workflow, the updated HPO term categorizations, flagged diseases, clinician comments and the re-ranked shortlist of variants are returned to the diagnostic laboratory for interpretation.

### Variant prioritization and presentation of HPO terms

We illustrate the variant prioritization steps in **Figure S2**. The VCF and starting HPO information are used for an Exomiser^14^ analysis. Exomiser is a gene prioritization tool that combines a score quantifying the likely pathogenicity of a variant associated with a gene (variant score) with a second score that measures the similarity of the phenotypic features of the patient with those associated with a pathogenic variant of the gene (phenotype score). Both values are used to generate the combined score for the variant.

We describe the prioritization and filtering steps in detail in the **Supplementary Information**. An Exomiser run is initiated based on the patient’s VCF and the HPO terms from the test requisition. This generates a list of variants, each of which is associated with a gene and one or more diseases, which in turn are associated with HPO terms present in individuals who have pathogenic variants of that gene, all of which are collected at his point. We then filter HPO terms for redundancy, focusing on those that are descendants of ‘Phenotypic abnormality’ (HP:0000118), and those associated with the specific disease prioritized by Exomiser. This leaves a subset of HPO terms that go through multiple rounds of selection, with the goal to present to the clinician ≤5 non-redundant HPO terms associated with each candidate gene, generating a maximum of 25 new HPO terms for categorization.

### Re-analysis based on enriched HPO information and disease exploration

A further potential input by the clinician is made possible by re-running Exomiser, now updated with the newly categorized HPO terms. The modified list of HPO terms updates the Exomiser phenotype to generate revised combined scores. These allow the refinement of the findings for clinician review, listed by gene, presenting the new list of genes ranked by the absolute combined score, displaying the magnitude and direction of change of the scores, and linking to associated candidate diseases. The hyperlink embedded with the names of each candidate disease brings the clinician to a description of the disease, letting them judge whether the syndrome of features plausibly fits their patient. Any disease of interest can be flagged, with the final step of the dive involving the clinician returning the list of updated HPO terms, any flagged diseases and free text comments as a file for the diagnostic laboratory.

### Software system architecture

We provide a detailed description of the GenomeDiver software in the **Supplementary Information** section.

## RESULTS

### GenomeDiver performance with simulated data

A video of the diagnostic laboratory interaction with the interface to set up a patient in the GenomeDiver system appears as **Supplementary Video 1**. Once set up, this allows a clinician to access the separate interface shown in **Figure 1** for categorization of HPO terms. The upper section ‘Add Phenotype Feature’ allows text to be entered, prompting HPO terms as a dropdown list for selection by clicking. The design is intended to focus the clinician on the middle section to ‘Classify All Phenotype Features’, dragging and dropping individual HPO terms into categories of ‘Present’, ‘Absent’ or ‘Unknown’ (not known whether present in the patient). After all terms have been categorized, the ‘Submit’ button on the bottom right activates and the updated information is sent back to GenomeDiver. This interaction appears in **Supplementary Video 2**.

**FIGURE 1:**
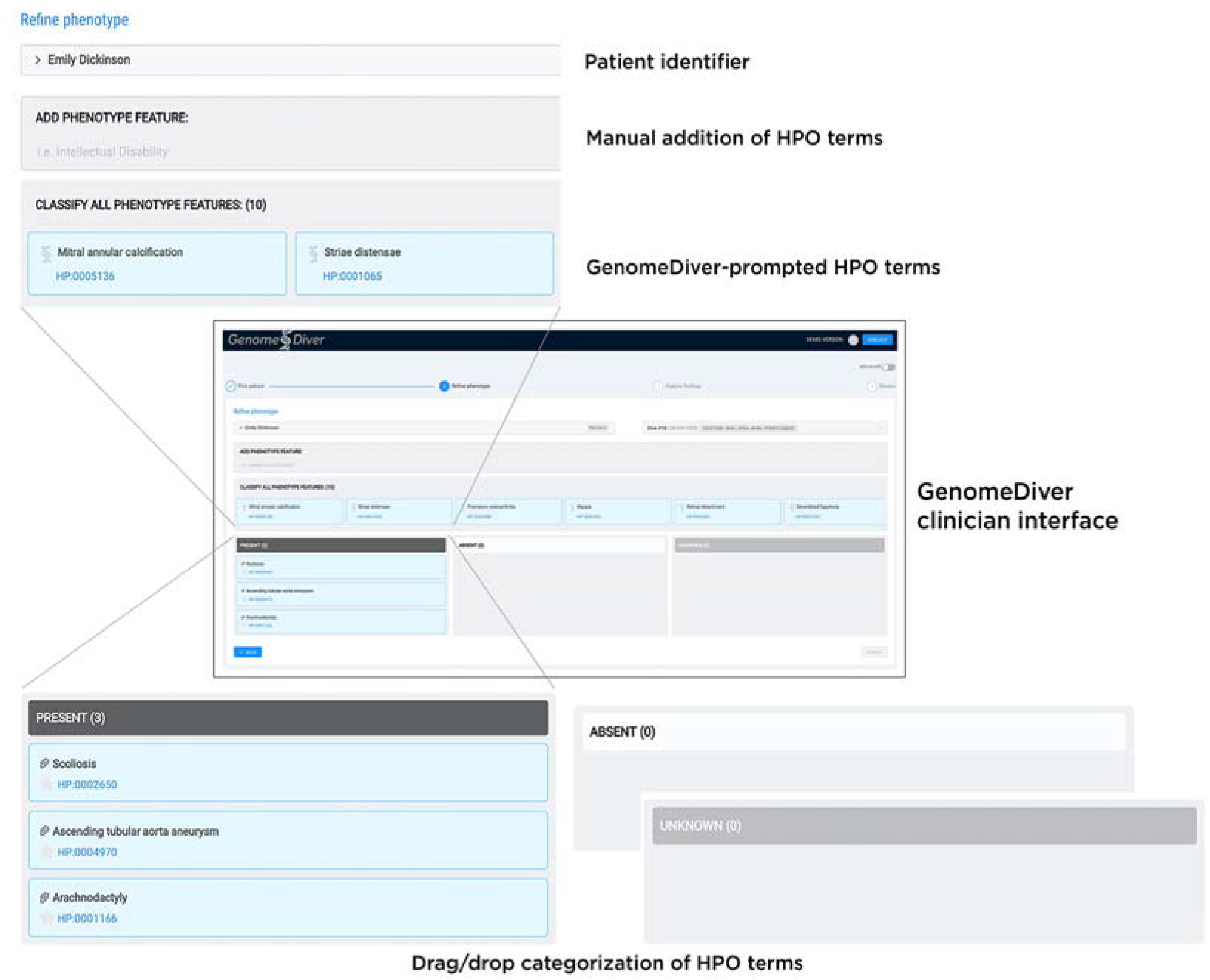
The clinician interface for GenomeDiver. HPO terms can be added in the ‘Add Phenotype Feature’ field by entering text which will bring up a list of HPO terms from which the appropriate term can be chosen. The HPO terms generated by GenomeDiver are shown in the ‘Classify all phenotype features’ section. These can be dragged into each of the categories underneath, ‘Present’, ‘Absent’ or ‘Unknown’. Three terms have been moved into the ‘Present’ category as an example.

We use the VCF for the publicly-available NA12878 genome,^15^ adding a variant of uncertain significance in the *FBN1* gene (ClinVar accession VCV000200085.4, NM_000138.4(FBN1): c.6449G>T(p.Arg2150Leu)). To simulate a request for analysis of a patient presenting with a clinical suspicion of Marfan syndrome (OMIM 154700), we used HPO terms for ascending tubular aorta aneurysm (HP:0004970), scoliosis (HP:0002650) and arachnodactyly (HP:0001166) as those potentially used in a test requisition in a clinical scenario.

We detail each step of the process of variant and gene filtering and prioritization in the **Supplementary Information** section (**Figure S2**), as well as the selection of HPO terms for presentation to the clinician (**Figures S3-S4**). For this particular combination of genomic and phenotypic information, two genes were selected, *FBN1* and *SKI*, the latter implicated in Shprintzen-Goldberg syndrome (OMIM 182212), whose phenotypic features likewise include aortic aneurysm, scoliosis and arachnodactyly.^16^

The HPO terms presented to the clinician are listed in **Supplementary Table 1**. In our experience, the process of categorization of ≤25 HPO terms in practice takes the user no more than ∼3 minutes, once reasonably familiar with the interface. We show how each term in our sample case was categorized in **Supplementary Table 1** and illustrate how a second Exomiser run presents candidate diseases in **Figure 2** (using steps illustrated in **Figure S5**). Of note, GenomeDiver never exposes information about variants to the clinician, preventing the clinician from bypassing the formal laboratory diagnostic reporting process.

**FIGURE 2:**
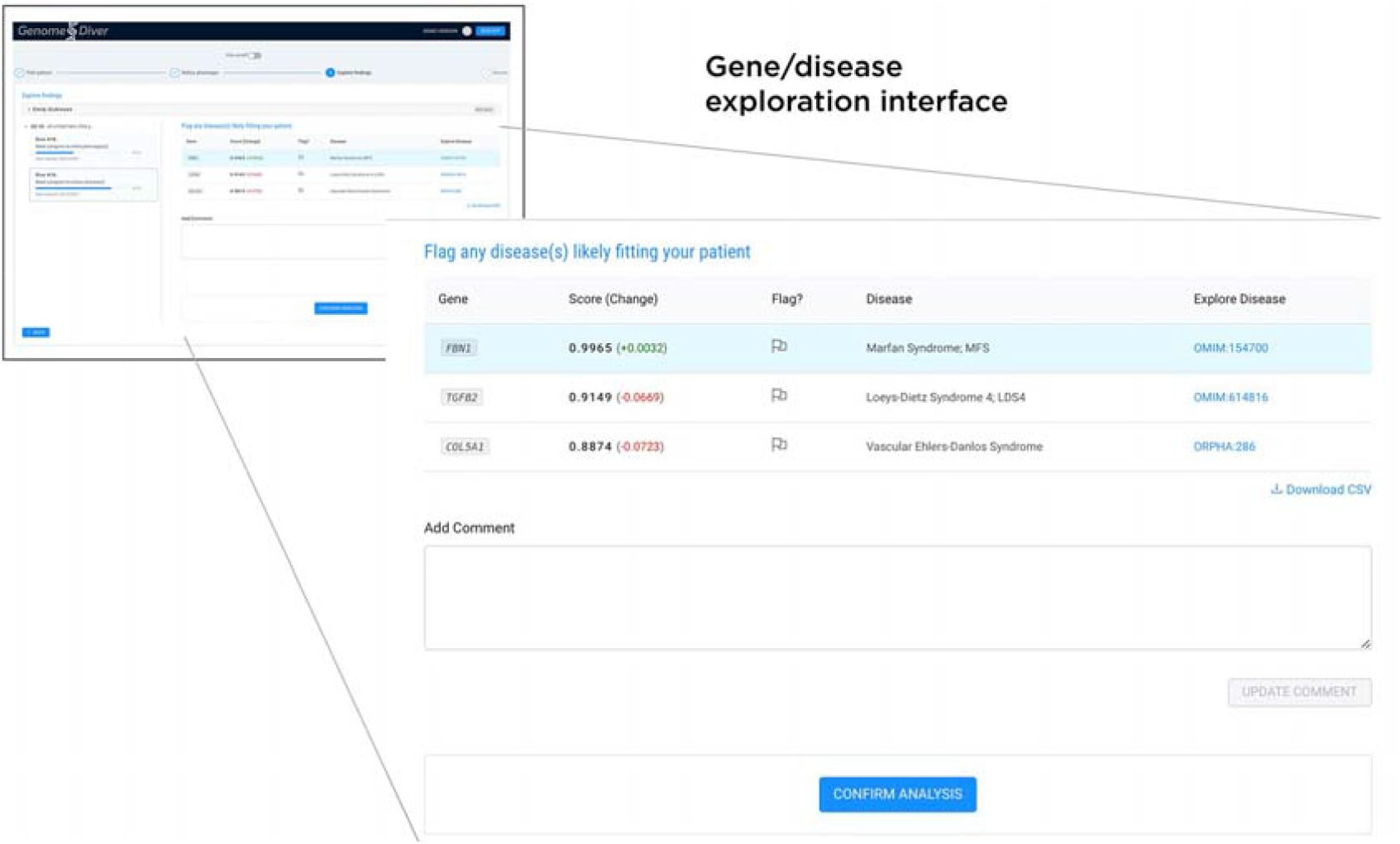
Following the categorization of the HPO terms, a second Exomiser run re-prioritizes variants, genes and associated diseases. The clinician is presented with a shortlist of genes, ranked by the Exomiser combined_score which is shown, as well as the change in score (green positive, red negative) resulting from the HPO term categorization. We list the disease and a link that allows the clinician to explore whether a disease description resembles their patient, allowing them to flag one or more as being candidates for causing the patient’s presentation. Free text can be used in the ‘Add comment’ box, finalizing the process by clicking ‘Confirm analysis’ to return the information to the diagnostic laboratory.

## DISCUSSION

In this initial application of GenomeDiver, we focused on improving the diagnostic process when performing genome-wide (exome, genome) sequencing in the diagnosis of rare diseases. We recognize that we can potentially improve both the rate of successful diagnosis and decrease the time spent by diagnostic laboratory personnel if we can facilitate the provision of the HPO terms that are most likely to discriminate the highest ranked gene candidates for causing the patient’s disease. With the appreciation that clinician time is limited, the interface is designed to maximize intuitiveness and ease of use, limiting the number of candidate HPO terms so that the entire categorization session should last no more than several minutes per patient. The second stage of input, facilitating the exploration of diseases that could be affecting the patient, permits further clinician insights to be provided, this time studying the syndrome of phenotypic features in candidate diseases. These steps of identifying individual phenotypic features and then considering how they might aggregate in diseases and syndromes reflect reasonably accurately the typical clinical genetics evaluation, but with the added intent of contributing to the laboratory diagnostic process. While we present certain choices for variant and HPO prioritization in the current version, these are intended to serve as the basis for an initial, functional version of GenomeDiver, but with the goal of incorporating community input for algorithmic improvement over time.

It is likely that HPO term harvesting from sources like EHRs^17^ and image analysis^18^ represents an area of innovation that will expand over time. A value of the GenomeDiver interface is that it allows a clinician to assess whether candidate terms gleaned from diverse, automated sources are actually present in a patient, creating the potential that GenomeDiver can act as a clearinghouse for downstream curation of candidate phenotypic features. The categorization of HPO terms as ‘absent’ is not yet used by Exomiser, but the updated LIRICAL approach, based on a likelihood ratio framework that includes information about HPO prevalence data,^19^ can exploit such information and will be served by this functionality within GenomeDiver.

There are more potential uses for GenomeDiver than for the initial diagnosis of a patient with a rare disease. GenomeDiver can facilitate re-analysis of patients with initially uninformative results whose phenotypes may have changed over time, and whose genomic variants may have undergone re-classification. GenomeDiver is fundamentally a tool to prompt ‘reverse phenotyping’ – the evaluation whether specific phenotypic features are present in a patient based on their genotypic information. As such, GenomeDiver has potential to serve clinicians not trained in clinical genetics and dysmorphology. With the development of HPO terms that now also encompass common diseases,^20^ GenomeDiver’s potential value as a reverse phenotyping tool that generates a genotype-based differential diagnosis could accordingly extend to much of medical genomics.

## Supporting information

Supplementary Methods and Results

Video S1 (compressed)

Video S2 (compressed)

## Data Availability

The manuscript describes software but is not reporting data. The software is available at https://github.com/GenomeDiver/

## DATA AVAILABILITY

The GenomeDiver software is available at https://github.com/GenomeDiver/,

## ACKNOWLEDGMENTS

Research reported in this publication was part of the NYCKidSeq project, supported by the National Human Genome Research Institute and National Institute for Minority Heath and Health Disparities of the National Institutes of Health under Award Number 1U01HG0096108.

## AUTHOR INFORMATION

Conceptualization NP CS KS FB NAH KB JAO SAS BDG EEK VJ TB JMG, Data curation VJ, Formal analysis NP FB CG TB JMG, Funding acquisition CRH MW BDG EEK, Investigation NP CS KS FB TB JMG, Methodology NP CS KS FB CG VJ TB JMG, Project administration NP FB GB EEK CG VJ TB JMG, Resources CRH MW BDG EEK CG VJ TB JMG, Software NP CS KS FB JMG, Supervision GB EEK CG JMG, Validation NP CS KS FB, Visualization NP CS KS FB JMG, Writing – original draft NP CS KS CG JMG, Writing – Review & Editing NP CS KS FB NAH GB KB JAO SAS CRH MW BDG EEK CG TB JMG.

