## Supplementary Methods and Results for "GenomeDiver: A platform for phenotype-guided medical genomic diagnosis"

**SUPPLEMENTARY INFORMATION**

**Supplementary Methods**

We illustrate the overall GenomeDiver workflow in terms of the user experience in **Figure S1**.

There are two major steps involved in selecting the set of Human Phenotype Ontology (HPO) terms that ends up being presented to the clinician for categorisation. The first uses the variant call format (VCF) file to identify variants that could be damaging, and performs filtering and annotation to generate a list of genes with sequence variants that are potentially causing disease.

The second step takes the HPO terms associated with these genes and performs filtering and prioritization, followed by a selection process that is intended to select those terms that are most likely to discriminate between the candidate genes.

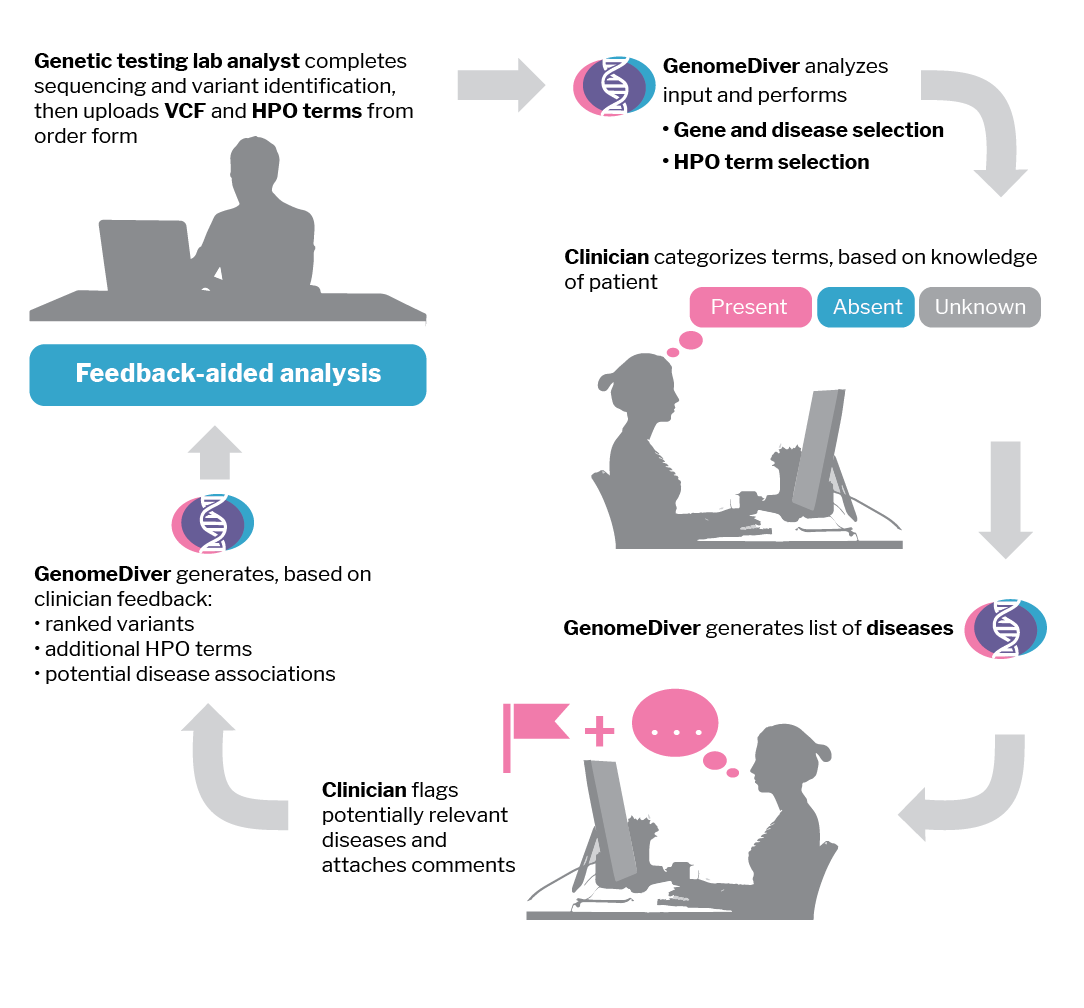

**Figure S1:** Overview of the GenomeDiver workflow, from the perspective of the diagnostic laboratory and clinician users.

**Gene and variant selection**

The software diagram for these steps is illustrated in **Figure S2**.

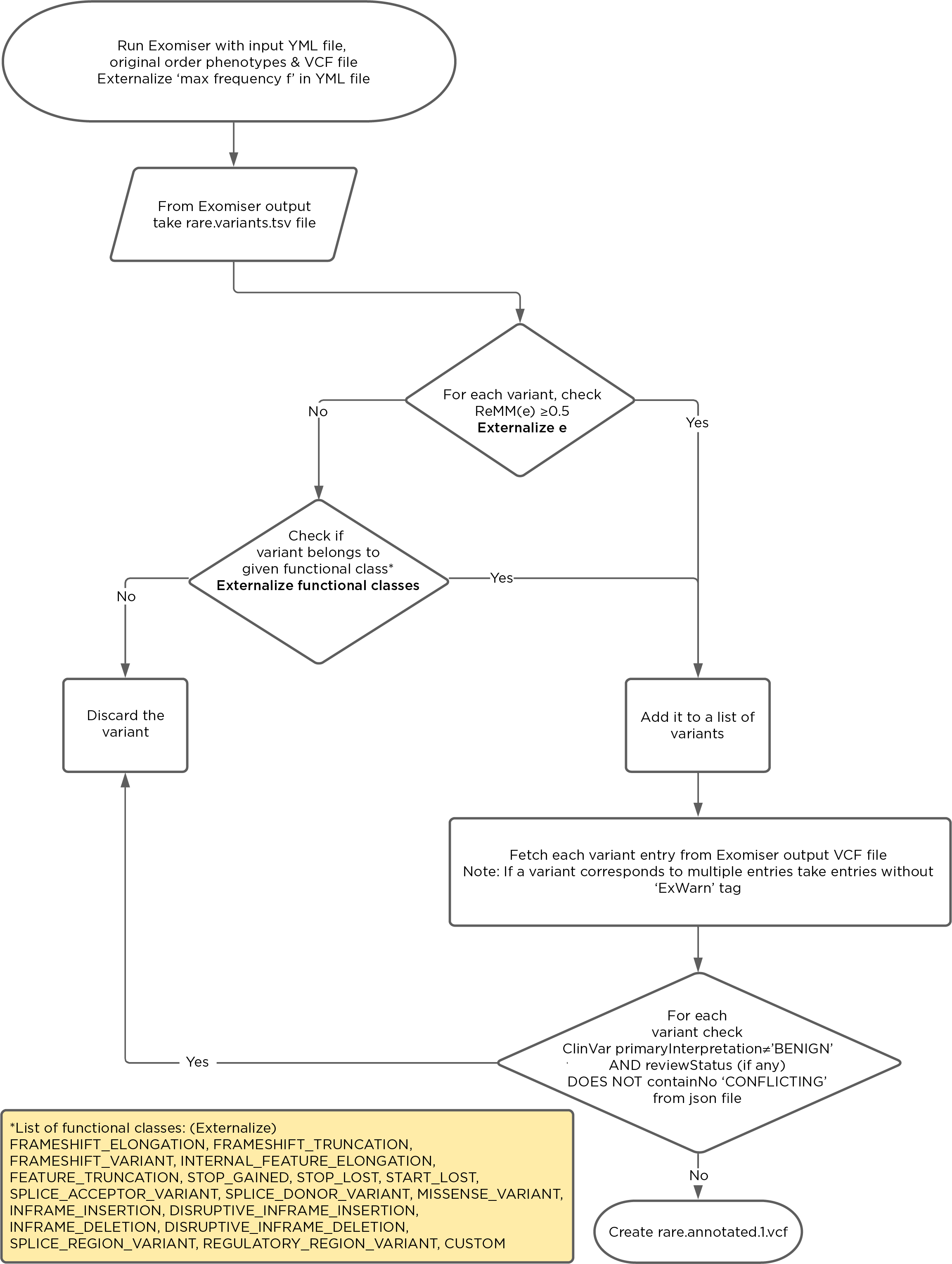

**Figure S2:** Software diagram of the gene and variant selection in steps 1-4.

**Step 1:** Exomiser is run using the patient’s VCF file and the HPO terms supplied with (or determined from) the test requisition. We use settings from the *exomiser.yml* file. At this stage, we accept all variants that do not exceed 10% allele frequency in any of the reference populations used by Exomiser.

Files generated: *rare.vcf*, *rare.variants.tsv* and *rare.genes.tsv*, the standard Exomiser output.

**Step 2:** We sort the *rare.vcf* file by genomic location using Picard:

http://broadinstitute.github.io/picard/

File generated: *rare.sorted.vcf*

**Step 3:** We annotate this file using VCFanno^2^ to add ClinVar information about the variants.

**Step 4:** We filter the variants using three criteria:

a. ClinVar clinical significance (CLINSIG), selecting anything categorized using the term ‘pathogenic’ and discarding all with ‘benign’ unless also described as conflicting.

b. For the remaining variants, we filter to select variants with allele frequencies all <2% in the reference populations, and regulatory Mendelian mutation (ReMM)^3^ scores >0.5

c. We retain variants with the following functional classes:

'FRAMESHIFT_ELONGATION', 'FRAMESHIFT_TRUNCATION', 'FRAMESHIFT_VARIANT', 'INTERNAL_FEATURE_ELONGATION', 'FEATURE_TRUNCATION', 'STOP_GAINED', 'STOP_LOST', 'START_LOST', 'SPLICE_ACCEPTOR_VARIANT', 'SPLICE_DONOR_VARIANT', 'MISSENSE_VARIANT', 'INFRAME_INSERTION', 'DISRUPTIVE_INFRAME_INSERTION', 'INFRAME_DELETION', 'DISRUPTIVE_INFRAME_DELETION', 'SPLICE_REGION_VARIANT', 'REGULATORY_REGION_VARIANT', 'CUSTOM'.

File generated: *rare.annotated.1.vcf*

**HPO term selection**

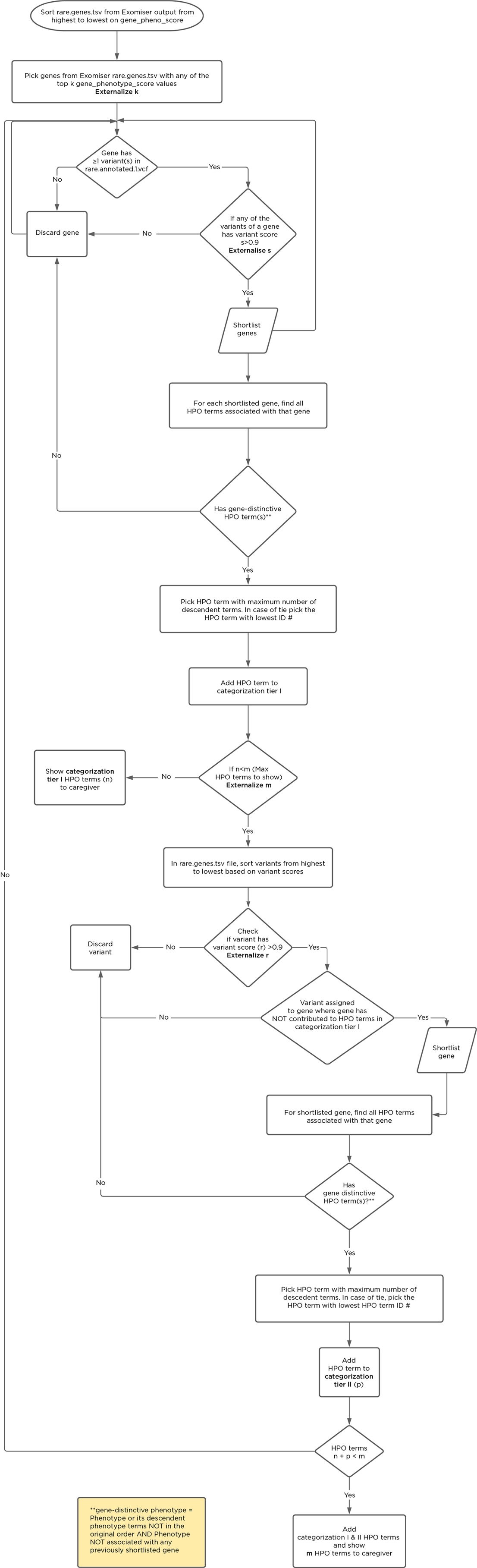

**Figure S3:** Software diagram of the HPO term selection in steps 5-13.

The software diagram for these steps is illustrated in **Figure S3**.

**Step 5:** From *rare.genes.tsv*, choose the genes with the top 10 gene_pheno scores from Exomiser. Retain only those genes with a variant listed in *rare.annotated.1.vcf*.

**Step 6:** Extract every HPO term occurring in all the diseases associated with each gene.

**Step 7:** Identify and retain HPO terms used in the original Exomiser run above. Filter out any related HPO terms that occur at a higher level on the HPO hierarchy as they are redundant and contain less information.

**Step 8:** Remove any terms that are not descended from HP:0000118 Phenotypic abnormality.

**Step 9:** Exomiser will have generated (in *rare.genes.tsv*) a predicted disease for each gene with a damaging variant. The HPO terms specifically occurring in that disease are now selected.

**Step 10:** The top 10 genes with variant_score values ≥0.90 are assigned to a Category 1, the top 10 of those remaining with variant_score values ≥0.85 are assigned to Category 2.

**Step 11:** Genes are now excluded unless they have a potentially damaging variant and are found in *rare.annotated.1.vcf*.

**Step 12:** Using the Phenotype Ontology Library (Phenol, https://phenol.readthedocs.io/), the HPO terms for each gene are ranked by their frequency of occurrence in the disease.

**Step 13:** The genes in Category 1 are ranked by pheno_score. The top gene selects the HPO term with the highest frequency value (if tied, a random choice is made). Each ranked gene sequentially gets to choose its top frequency HPO term. After one round of selections, the genes are re-ranked by variant_score and the process is repeated up to 5 rounds total or until no more HPO terms can be selected. The maximum number of HPO terms that can be selected by a gene is 5. The goal is to reach 25 HPO terms in total, which may require re-initiating the process in the Category 2 genes.

This list of ≤25 HPO terms is then presented to the clinician in the GenomeDiver user interface.

**Example: NA12878 genome with *FBN1* variant of uncertain significance**

We created an artificial human genome using the public NA12878 genome^4^ into which we added a sequence change classified in ClinVar as a variant of uncertain significance (VUS) in the *FBN1* gene (ClinVar accession VCV000200085.4, NM_000138.4(FBN1): c.6449G>T (p.Arg2150Leu)).

To initiate Step 1 above, we used this VCF file and three HPO terms (Ascending tubular aorta aneurysm HP:0004970, Scoliosis HP:0002650 and Arachnodactyly HP:0001166), representing the kind of information that might be submitted for a patient with suspected Marfan syndrome. At the end of Step 4, the resulting *rare.annotated.1.vcf* file included 17,275 variants.

Step 5 results, identifying the top 10 genes from *rare.genes.tsv* ranked by gene_pheno scores generated the following list:

*FBLN5*

*FBN1*

*TGFBR1*

*TGFBR2*

*ZMPSTE24*

*SKI*

*SLC2A10*

*TGFB2*

*MYLK*

*PRKG1*

Of this 10, only *FBN1* and *SKI* had variants in *rare.annotated.1.vcf* (Step 11). *FBN1* had a variant_score of 1.000, and was assigned to Category 1, with *SKI* assigned to Category 2 based on its variant_score of 0.8823 (Step 10). As these are the genes that emerged from these analyses, we will focus on what happened with them specifically.

The number of HPO terms for the next several steps are illustrated in **Figure S4**. While Step 7 removed the three input HPO terms for both genes, for *FBN1* the term Aortic aneurysm HP:0004942 was also removed as it exists higher on the hierarchical tree than Ascending tubular aorta aneurysm HP:0004970. Step 8 removed Autosomal dominant inheritance HP:0000006 for both genes but also Sporadic HP:0003745 for *SKI*. Steps 9-11 remove the majority of HPO terms, with the final selection of 5 HPO terms occurring in Step 13.

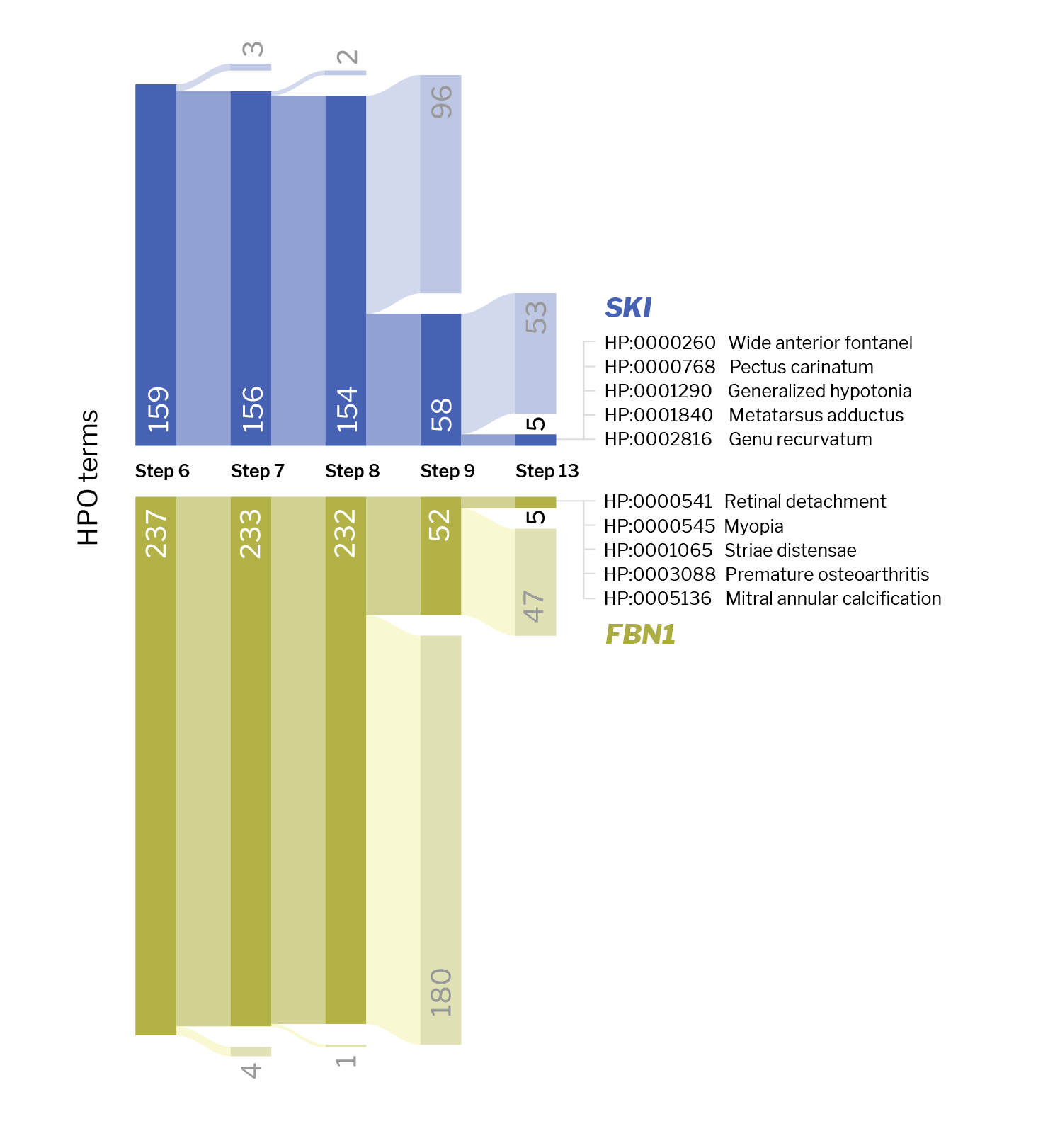

**Figure S4:** The number of HPO terms at each step of the selection process is illustrated for each gene in our NA12878/*FBN1* example, culminating in the selection of 5 terms per gene.

The software diagram illustrating the presentation of genes and diseases to the clinician is shown in **Figure S5**.

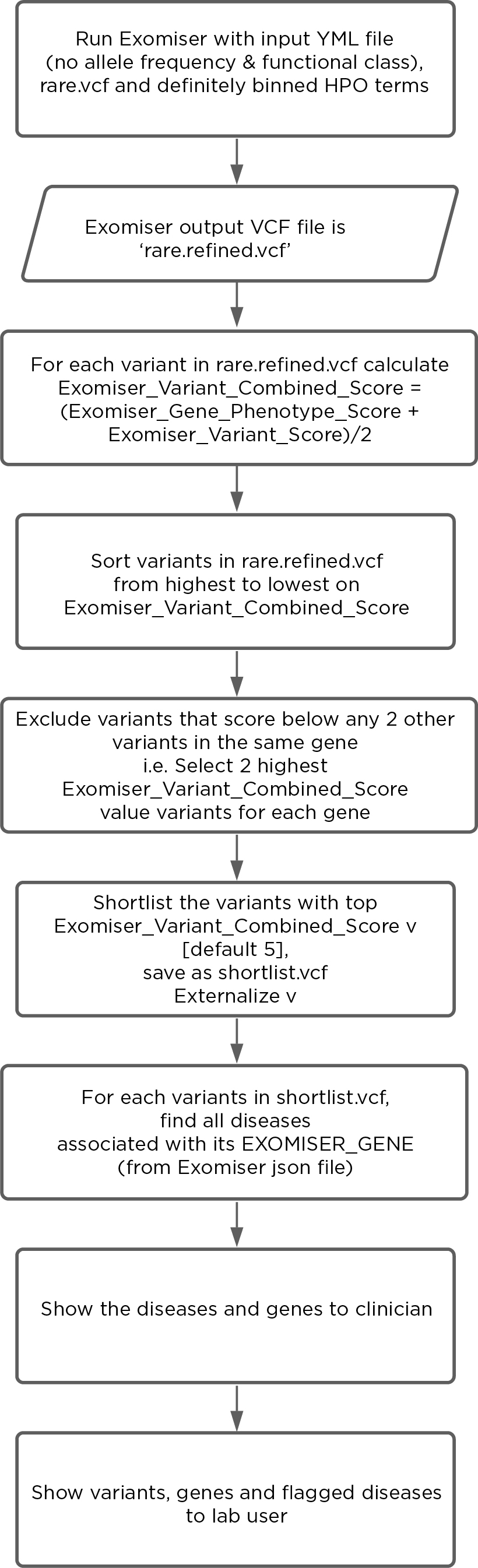

**Figure S5:** Software diagram showing how the genes and diseases are selected for presentation to the clinician.

**Software system architecture**

GenomeDiver is written in the Scala programming language, which uses terse code and prioritizes concurrency, the techniques and mechanisms that enable a computer program to perform multiple different tasks simultaneously, essential for scalability. Scala’s targeting of the Java Virtual Machine (JVM) allows GenomeDiver to be natively compatible with the sequencing tools developed by the Broad Institute, such as GATK, Picard and HTSJDK, and HPO tools developed in Java.

The chosen Scala frameworks (Akka, Sangria) on top of the language provide the backbone of application to operate under FAIR guiding principles^5^ at the scale of genomic data and the web security standards required for the patient data in GenomeDiver. Sangria (GraphQL implementation) provides efficient interoperability to other systems using the schema-driven GraphQL query language, effectively forming a robust and standard adapter layer for application programming interfaces (APIs).

All analyses are carried out using the Nextflow bioinformatic workflow manager. Nextflow provides reproducibility and reusability of computational pipelines through a domain specific language. This allows the underlying analysis pipelines to be easily extracted and verified, independent of GenomeDiver.

GenomeDiver is currently divided into three main parts: the application, its database, and a supporting workflow runner. The build and deployment are currently being managed by Node and Sbt, and the built application is then deployed to a virtual machine that submits to an internal Slurm cluster.

For the front end, GenomeDiver uses React, a re-usable JavaScript library of components with standardized markup. The Nextflow bioinformatic workflow manager runs the bioinformatic pipelines. The pipelines run tools such as Exomiser^1^ and VCFAnno.^2^ GenomeDiver’s own genomic/phenotype filters use Htsjdk and Phenol. Nextflow is used to schedule and run analysis tasks (Exomiser) on a cluster manager. The HPO ontology is made searchable by the web interface, and the HPO annotations are used for prioritizing phenotypes.

GenomeDiver is designed to be accessible to as many potential users as possible. Its development is guided by the W3C Web Content Accessibility Guidelines (WCAG) 2.1 from 2018 (www.w3.org/TR/WCAG21/). Currently, front end color profiles are tested using contrastchecker.com to ensure that all those with color vision deficiency are able to discriminate the color palette used.

**Supplementary Tables**

**Supplementary Table 1**

We show the HPO terms presented in our NA12878/*FBN1* example and how they were categorised to generate the results in **Figure 2**.

| **HPO term** | **HPO identifier** | **Gene association** | **Categorization** |
| --- | --- | --- | --- |
| Ascending tubular aorta aneurysm | HP:0004970 | *FBN1 SKI* | Present (input) |
| Scoliosis | HP:0002650 | *FBN1 SKI* | Present (input) |
| Arachnodactyly | HP:0001166 | *FBN1 SKI* | Present (input) |
| Mitral annular calcification | HP:0005136 | *FBN1* | Present |
| Striae distensae | HP:0001065 | *FBN1* | Present |
| Premature osteoarthritis | HP:0003088 | *FBN1* | Present |
| Myopia | HP:0000545 | *FBN1* | Present |
| Retinal detachment | HP:0000541 | *FBN1* | Present |
| Pectus carinatum | HP:0000768 | *FBN1 SKI* | Present |
| Metatarsus adductus | HP:0001840 | *SKI* | Absent |
| Genu recurvatum | HP:0000047 | *SKI* | Absent |
| Generalised hypotonia | HP:0001290 | *SKI* | Absent |
| Wide anterior fontanel | HP:0000260 | *SKI* | Absent |

**Supplementary Videos**

**Supplementary Video 1**

A video of the diagnostic laboratory interaction with the interface to set up a patient in the GenomeDiver system is shown.

**Supplementary Video 2**

We show an example of the interface for the clinician.
